# Electronic Genome Mapping Enables High-Resolution Cytogenomic Profiling of Hematologic Malignancies: A Proof-of-Principle Study

**DOI:** 10.64898/2026.09.11.26362883

**Authors:** Ashis K. Mondal, Vishakha Vashisht, Yang Zhang, Michael D. Gallagher, Shuk Shukor, Michael Diaz, Ashutosh Vashisht, Alka Chaubey, Ravindra Kolhe

## Abstract

**Background/Objectives:** Many myeloid malignancies are defined by recurrent structural variants (SVs) and copy-number alterations (CNAs) under the current World Health Organization (WHO) and International Consensus Classification (ICC) frameworks. Detecting these abnormalities typically requires several standard-of-care (SOC) assays, each with its own blind spots, and then laboratories and clinicians are left to piece the results together. Genome mapping has emerged as a mainstream technology with the potential to serve as a high-resolution alternative to several standard-of-care cytogenetic assays such as karyotyping (KT), fluorescence in situ hybridization (FISH) or chromosomal microarray (CMA). Our laboratory at Augusta University has published numerous publications on the analytical and clinical utility of optical genome mapping (OGM) as a first-line assay for hematological malignancies. Electronic genome mapping (EGM) pursues the same goal with an electronic rather than optical readout. The objective of this study was to demonstrate the performance and utility of EGM as a valuable tool to detect clinically relevant SVs and CNAs found by SOC testing. Cytogenomic characterization of hematologic malignancies commonly requires multiple complementary assays, including karyotyping, FISH, and chromosomal microarray, each with distinct analytical limitations. Electronic genome mapping (EGM) provides genome-wide interrogation of structural variants and copy-number alterations using electronic single-molecule detection. We evaluated the feasibility and analytical performance of EGM in previously characterized myeloid malignancies.

**Methods:** EGM was performed on the Nabsys OhmX platform using five previously characterized bone marrow aspirates from cases with acute myeloid leukemia (AML) or chronic myeloid leukemia (CML). EGM calls were compared with findings from karyotyping (KT), fluorescence in situ hybridization (FISH), and OGM.

**Results:** All five samples passed quality control, with contig N50 values ranging from 1.8 to 3.8 Mb and more than 90% of reference labels represented in the assembled contigs. EGM detected every SOC-reported abnormality including two t(9;22)(q34.12;q11.23)/*BCR::ABL1* rearrangements, trisomy 4, trisomy 21, and monosomy 7. In the most complex case, EGM resolved a del(20q) reported by KT and FISH into an intrachromosomal fusion, fus(20;20)(q11.21;q13.12), showing not only what was lost but where the chromosome rejoined. Like OGM, EGM identified an approximately 1.8 Mb loss at 7q22.1 involving 45 of genes of which only *CUX1* is associated with cancer. This 7q loss was not detected by KT or FISH. Interestingly, EGM also detected the *KMT2A-PTD* variant previously detected by OGM. Collectively, EGM demonstrated 100% concordance (6/6 SOC-reported abnormalities; 9/9 OGM calls).

**Conclusions:** In this pilot study on five hematological malignancy specimens, EGM matched SOC and OGM across translocations, aneuploidies, insertions and deletions, including *BCR::ABL1* rearrangements in both AML and CML, and added high-resolution breakpoint detail beyond karyotype resolution. These results support larger studies across more subtypes to establish EGM as a first-line genome-wide cytogenomic tool for hematological malignancies.

## 1. Introduction

The World Health Organization (WHO) classification of hematological neoplasms encompasses myeloid and lymphoid malignancies, many of which are defined or further characterized by recurrent genetic/genomic abnormalities.^1,2^ A comprehensive evaluation of these blood cancers relies on a combination of molecular and cytogenetic approaches to identify sequence variants, copy number abnormalities (CNAs), and structural variants (SVs).^2-5^ Advances in next-generation sequencing (NGS) over the past decade have substantially expanded the ability to characterize sequence-level alterations, with targeted gene panels and whole-exome sequencing increasingly replacing single-gene or single-variant testing.^5^

Since 1974, cytogenetic abnormalities remain central to the diagnosis and classification of many hematological malignancies and provide important prognostic and therapeutic information.^1-7^ Conventional cytogenetic testing typically relies on a combination of karyotyping (KT), fluorescence in situ hybridization (FISH), and chromosomal microarray (CMA) analysis, with each method providing a distinct but incomplete view of genome structure. Karyotyping offers a genome-wide assessment of chromosomal abnormalities but is limited by relatively low resolution. FISH provides higher-resolution interrogation of specific loci but is inherently targeted and generally requires prior knowledge of the genomic abnormality. Chromosomal microarrays provide substantially greater resolution for CNAs but cannot detect balanced rearrangements and provide limited information regarding the context or orientation of the complex genomic alterations.

These limitations are particularly relevant in hematologic malignancies where acquired or somatic structural rearrangements are common and may have direct diagnostic, prognostic, or therapeutic implications.^1-7^ Balanced translocations, for example, could potentially generate clinically important gene fusions that cannot be resolved at the gene level by KT. Similarly, submicroscopic duplications are detected as copy number gains without resolving their genomic insertion site or orientation, obscuring the underlying mechanism of the structural event. Complex rearrangements are more challenging because individual abnormalities may be detected by different assays without providing a complete picture of the resulting abnormal chromosomal architecture.

A comprehensive approach to cytogenomic analysis would therefore ideally combine genome-wide detection with sufficient resolution to identify and characterize a broad range of SVs, including balanced and unbalanced rearrangements, inversions, insertions, duplications, and complex genomic rearrangements. Genome sequencing (GS) has been investigated as a potential comprehensive approach; however, its clinical implementation remains associated with substantial bioinformatic requirements and challenges in the detection and interpretation of certain classes of SVs. In hematological malignancies, sequencing-based approaches have also frequently focused on recurrent translocations and large copy number abnormalities, with limitations in assessing only targeted genes (sequence) variants and inability to discern structural context of more complex genomic events.^4-11^ Thus, despite advances in sequencing technologies, there remains a need for genome-wide cytogenomic approaches capable of detecting and characterizing SVs at higher resolution while preserving the genome-wide perspective of conventional cytogenetics.

Genome mapping approaches have emerged as a means of interrogating structural variation and copy number change across the genome in a single analysis. Most published examples in hematologic malignancies have been generated using optical genome mapping (OGM)^9^. Multiple published studies have demonstrated feasibility, concordance, and implementation of OGM for the detection of recurrent translocations, aneuploidies, copy number alterations, and complex rearrangements in acute myeloid leukemia, myelodysplastic neoplasms, acute lymphoblastic leukemia, and other hematologic neoplasms^9-12^ . These studies establish the value of high-resolution genome mapping for examining abnormalities that are usually missed by conventional cytogenetic methods. Previously published studies from our laboratory provide a useful framework for evaluating new mapping technologies in well-characterized specimens without presuming that early technical concordance is equivalent to clinical validation ^7,11^. The ability to interrogate structural variation and copy number changes in a single genome-wide assay provides an opportunity to consolidate information that otherwise may require multiple complementary techniques. ^6-11^

Electronic genome mapping (EGM) is another long-read genome mapping technique that detects the positions of sequence-specific tags (labels) along individual high-molecular-weight (HMW) DNA molecules electronically. The resulting single-molecule reads can be assembled into contigs, directly aligned to a reference, and analyzed for changes in label spacing, molecule organization, and genomic representation, enabling detection of structural rearrangements and copy number abnormalities. EGM uses an electronic rather than imaging-based detection architecture, and therefore, it represents a distinct approach of genome mapping. However, its performance relative to conventional cytogenetic methods and OGM in hematological neoplasms has not yet been described in a peer-reviewed cohort.

In this proof of principle, pilot study, we evaluated the performance of EGM for the detection and characterization of clinically relevant or pathogenic genomic abnormalities in hematological malignancies and compared its findings with those obtained using current standard-of-care cytogenetic methods, including karyotyping, FISH, and OGM. The specimens were selected to include several SV classes including a recurrent translocation, whole-chromosome gains and losses, interstitial deletion, and a more complex structural rearrangement.

## 2. Methods

### 2.1 Specimens and Standard-of-Care (SOC) Results

Five bone marrow aspirate (BMA) specimens were obtained from cases with myeloid malignancies at the Georgia Esoteric Molecular laboratory at Augusta University and stored at −80°C until processing. The cohort included four acute myeloid leukemia (AML) specimens and one chronic myelogenous leukemia (CML) specimen. The samples used and processed in this study were collected under a study approved by the Institutional Review Board (IRB #00000150, HAC IRB #611298, 4 April 2018) at Augusta University, GA, USA. Standard-of-care testing (KT, FISH, and/or OGM) results were available for all five specimens. The reported KT, FISH, and OGM findings were used for establishing EGM performance and concordance. The primary endpoint was event-level concordance between EGM and previously reported SOC findings. Secondary analyses assessed concordance with OGM and additional structural characterization provided by EGM.

### 2.2 Electronic Genome Mapping

HMW DNA was extracted from BMA samples using the NEB Monarch HMW DNA Extraction Kit for Cells & Blood (T3050L, New England Biolabs, Inc, Ipswich, USA) according to the Nabsys DNA Isolation for OhmX™ Genome Preparation Guide (Nabsys 2.0 LLC, Providence, USA). Detailed protocols for HMW genomic DNA sample preparation and data acquisition from the OhmX Platform are available from Nabsys (https://www.nabsys.com/learn/sample-prep-kit). Briefly, the standard protocol begins with nicking of HMW DNA and incorporation of biotinylated dNTPs at the nick sites during a simultaneous nicking and labeling step. The labeled DNA is subsequently purified to remove unincorporated dNTPs and then tagged with a streptavidin-DNA complex. The tagged DNA is coated with protein to straighten and stiffen the HMW DNA molecules prior to analysis. The resulting DNA–protein complex is injected into the OhmX Analyzer, and data are collected at a coverage of 300x or greater. The resulting electrical signal pattern is converted into physical distances along individual DNA molecules automatically using High-Definition Mapping (HDM) Analysis software (v1.11.1302). Following signal processing, the resulting data are uploaded to Nabsys Navigator Software platform for data analysis and interpretation.

### 2.3 Data Analysis and Visualization

Structural variant analysis was performed using the Nabsys Navigator SV Discover pipeline (v1.0.0). These individual molecules are assembled into longer consensus contigs and aligned to the GRCh38 human reference label patterns. Differences in label patterns between contigs and reference alignments were identified as potential structural variants associated with hematological malignancy subtypes. General structural variant categories called by EGM are insertions, deletions, duplications, inversions, and translocations. The full SV list was annotated with QC information, masked region overlaps, and control database overlaps to allow specific filtering (e.g. false positive SVs, high-confidence SVs, SVs not in control databases). Candidate SV calls were reviewed using Nabsys Navigator chromosome-level displays, annotated SV list, and molecule-level evidence. Reported genomic coordinates and cytogenetic bands were based on GRCh38. The identified structural variants were compared at the event level against the corresponding karyotyping, FISH, and OGM results.

Copy number analysis was performed using the Nabsys CNV pipeline (v0.9.0). Reference-aligned molecules passing the pipeline’s two-dimensional quality threshold were counted in genomic bins after exclusion of masked regions. Bin counts were corrected using the pipeline’s negative-control reference, scaled, and log transformed. Segmentation was performed using the pruned exact linear time (PELT) algorithm, and genome-wide ploidy and copy number states were estimated at 1Mb bin sizes. Final structural variant and copy number results were tabulated for comparison with standard-of-care methods.

### 2.4 Data Quality Assessment

Data quality was assessed in Nabsys Navigator. Molecule-level metrics included estimated genome coverage per sample (>300x), molecule N50 (>65kb), and mean interval size (>4.0kb). Label density was assessed at approximately 22-25 labels per 100kb.

Assembly quality was assessed using the inferred chromosomal sex, contig N50, and percentage of reference sites covered (>80%). Inferred sex was compared with the reported sex for each specimen. Copy number profiles were additionally reviewed for genome-wide coverage uniformity and baseline stability. Sample-level data yield and quality metrics are summarized in Table 2.

## 3. Results

### 3.1 Cohort Characteristics and EGM analysis

The exploratory cohort comprised five previously characterized hematologic malignancy specimens, including four AML specimens and one CML specimen. The SOC findings represented several classes of chromosomal abnormalities: translocation, whole-chromosome gains and losses, copy number loss or deletion, and a complex intrachromosomal rearrangement. KT, FISH, and OGM results were available for all five specimens (Table 1). EGM identified the primary cytogenetic abnormality reported for each specimen and detected the corresponding OGM findings for these events.

**Table 1.** Comparison of KT, FISH, OGM, and EGM findings across the five hematologic malignancy specimens. Genomic coordinates are based on GRCh38. AML, acute myeloid leukemia; CML, chronic myeloid leukemia; EGM, electronic genome mapping; FISH, fluorescence in situ hybridization; KT, Karyotyping; OGM, optical genome mapping.

| Sample ID | Diagnosis | KT (ISCN) | FISH (notes) | OGM (ISCN) | EGM (egm) |
| --- | --- | --- | --- | --- | --- |
| AUG-664 | AML | 46,XY,del(20)(q13.1)[17]/46,XY[3] | Loss of chromosome 20q detected | ogm[GRCh38] ins(11;?)(q23.3;?)<br>ogm[GRCh38] 20q11.21q13.12 (32720656_45856309)x1~2<br>ogm[GRCh38] fus(20;20)(q11.21;q13.12)<br>ogm[GRCh38] 7q22.1 (100371063_102255480)x1~2 | egm[GRCh38] ins(11;?)(q23.3;?)<br>egm[GRCh38] 20q11.21q13.12x1~2<br>egm[GRCh38] fus(20;20)(q11.21;q13.12)<br>egm[GRCh38] 7q22.1(100386238_102250622)x1~2 |
| AUG-667 | AML | 45,XX,-7,t(9;22)(q34;q11.2)[17]/46,XX[3] | AML FISH revealed loss of chromosome 7 and *BCR::ABL1* fusion | ogm[GRCh38] 7p22.3q36.3x1~2<br>ogm[GRCh38] t(9;22)(q34.12;q11.23) | egm[GRCh38] 7p22.3q36.3x1~2<br>egm[GRCh38] t(9;22)(q34.12;q11.23) |
| AUG-668 | AML | 47,XY,+4 | Trisomy 4 | ogm[GRCh38] 4p16.3q35.2x2~3 | egm[GRCh38] 4p16.3q35.2x2~3 |
| AUG-679 | CML | 46,XX,t(9;22)(q34;q11.2)[20] | CML FISH was positive for *BCR::ABL1* fusion in 93.5% of cells | ogm[GRCh38] t(9;22)(q34.12;q11.23) | egm[GRCh38] t(9;22)(q34.12;q11.23) |
| AUG-687 | AML | 47,XY,+21 | Gain of *RUNX1* gene on 21 chr | ogm[GRCh38] 21q21.3q22.3 (28548156_42643859)x2~3 | egm[GRCh38] 21q21.3q22.3x2~3 |

### 3.2 Pre-secondary Analysis and Post-secondary Analysis Quality Control (QC)

Before secondary analysis, molecule-level metrics for each of the five specimens were tabulated from SVWB Molecule Stats Panel and HDMA Whole Genome Stats (Table 2). All five specimens met the recommended criteria for molecules N50, mean interval size, and label density. Estimated coverage exceeded 300x for four specimens, with AUG-668 at 292x, and label false-negative rate was below the recommended <11% for four specimens, with AUG-679 at 11.2% (data not shown). All specimens were advanced to secondary analysis. Molecules >65 kb with label density in the recommended range were used as input for the SV Discover assembly pipeline.

**Table 2.** Data QC summary for the hematological malignancy sample cohort. Recommended value cutoffs are provided in the bottom column.

| **Sample** | **Molecule N50 length (kb)** | **Contig N50 length (Mb)** | **Estimated Coverage** | **Label Density / 100 kb** | **% Contigs w/ 9+ labels** | **Covered sites ratio** |
| --- | --- | --- | --- | --- | --- | --- |
| **AUG-664** | 104.771 | 2.041 | 329.568 | 24 | 94.970 | 91.17% |
| **AUG-667** | 111.717 | 3.783 | 470.821 | 24 | 94.874 | 91.14% |
| **AUG-668** | 104.429 | 1.758 | 292.055 | 24 | 94.784 | 90.71% |
| **AUG-679** | 110.221 | 2.423 | 448.68 | 22 | 90.457 | 90.60% |
| **AUG-687** | 108.125 | 1.984 | 311.243 | 24 | 94.715 | 90.66% |
| **Recommended** | >65 kb | >0.5 Mb | >300X | 22 - 25 | >80% | >80% |

After secondary analysis, all five specimens met assembly-level QC criteria. Contig N50 ranged between 1.758 to 3.783 Mb, and covered sites ratio from 90.60% to 91.17%, well above the >80% minimum. Inferred chromosomal sex matched the reported sex for all five specimens.

### 3.3 Analytical Performance of EGM: 100% concordance with SOC results

#### 3.3.1 Detection of structural rearrangements and copy number abnormalities-100% concordant

EGM demonstrated complete concordance with the previously characterized abnormalities represented in this pilot cohort. EGM detected t(9;22)(q34.12;q11.23) in both specimens (AUG-667 and AUG-679) in which the rearrangement had been reported by KT, FISH, and OGM. AUG-667, an AML specimen with karyotyping results of 45,XX,-7,t(9;22)(q34;q11.2)[17]/46,XX[3], was a case example containing both structural and copy number abnormalities. EGM identified the t(9;22) rearrangement along with loss of chromosome 7, which was consistent with the KT, FISH and OGM results (Figure 1). In AUG-679, a CML specimen, FISH was positive for *BCR::ABL1* in 93.5% of cells and KT showed 46,XX,t(9;22)(q34;q11.2)[20]. Both OGM and EGM detected the rearrangement as t(9;22)(q34.12;q11.23) further refining the breakpoints reported by KT analysis (Figure 3).

**Figure 1.**
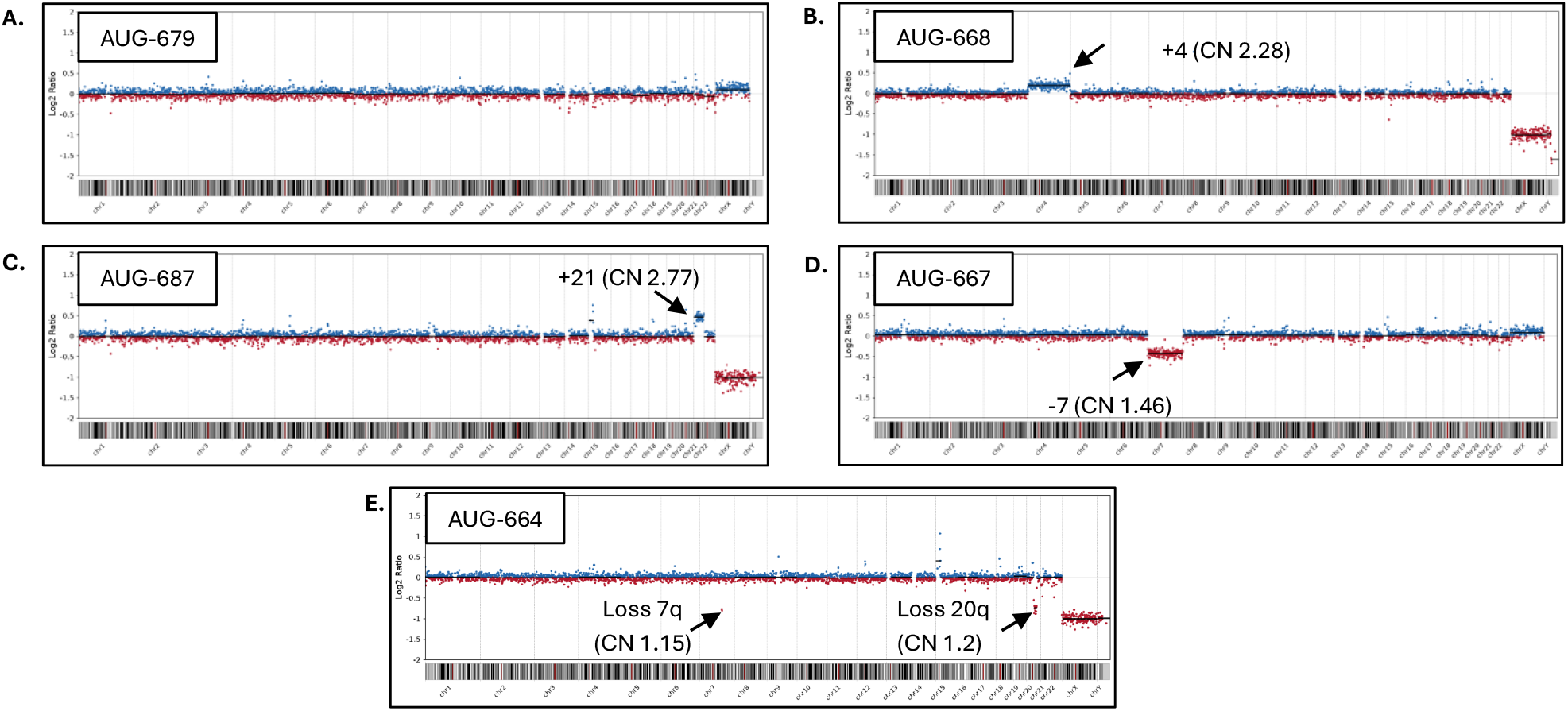
EGM copy number profiles across the cohort. **(A)** AUG-679, XX (female), no CNVs detected. **(B)** AUG-668, XY (male), with trisomy 4. **(C)** AUG-687, XY (male), with trisomy 21. **(D)** AUG-667, XX (female), with monosomy 7. **(E)** AUG-664, XY (male), with interstitial losses involving 7q and 20q. Genomic coordinates are based on GRCh38.

Whole-chromosome gains reported in the remaining AML specimens (AUG-668 and AUG-687) were also represented in both genome mapping results. EGM and OGM both identified chromosome 4 gain in AUG-668, consistent with trisomy 4 by FISH and KT. Chromosome 21 gain was detected by both EGM and OGM in AUG-687, consistent with gain of *RUNX1* material reported by FISH and trisomy 21 by KT. Together, these cases demonstrate EGM’s 100% concordance with SOC methods including KT, FISH, and OGM. Detailed comparisons among the four methods are provided in Table 1.

#### 3.3.2 EGM in a complex case (resolving the SV in a chromosome 20 abnormality)

AUG-664 contained the most structurally complex finding in the cohort. Chromosome analysis showed 46,XY,del(20)(q13.1)[17]/46,XY[3], and FISH identified an abnormal clone involving loss of chromosome 20 material. OGM and EGM each characterized this underlying chromosome 20 abnormality through complementary copy number and structural variant results. The copy number analysis identified the net loss of 20q material, while the structural variant analyses identified the junction as an intrachromosomal fusion, fus(20;20)(q11.21;q13.12) (Figure 2). The fusion therefore describes the deletion associated with the observed copy number loss. Both OGM and EGM also identified a copy number loss at 7q22.1 that was not reported by KT or FISH. Both OGM and EGM also identified a ∼8kb insertion within *KMT2A*, an important and actionable variant for AML.

**Figure 2.**
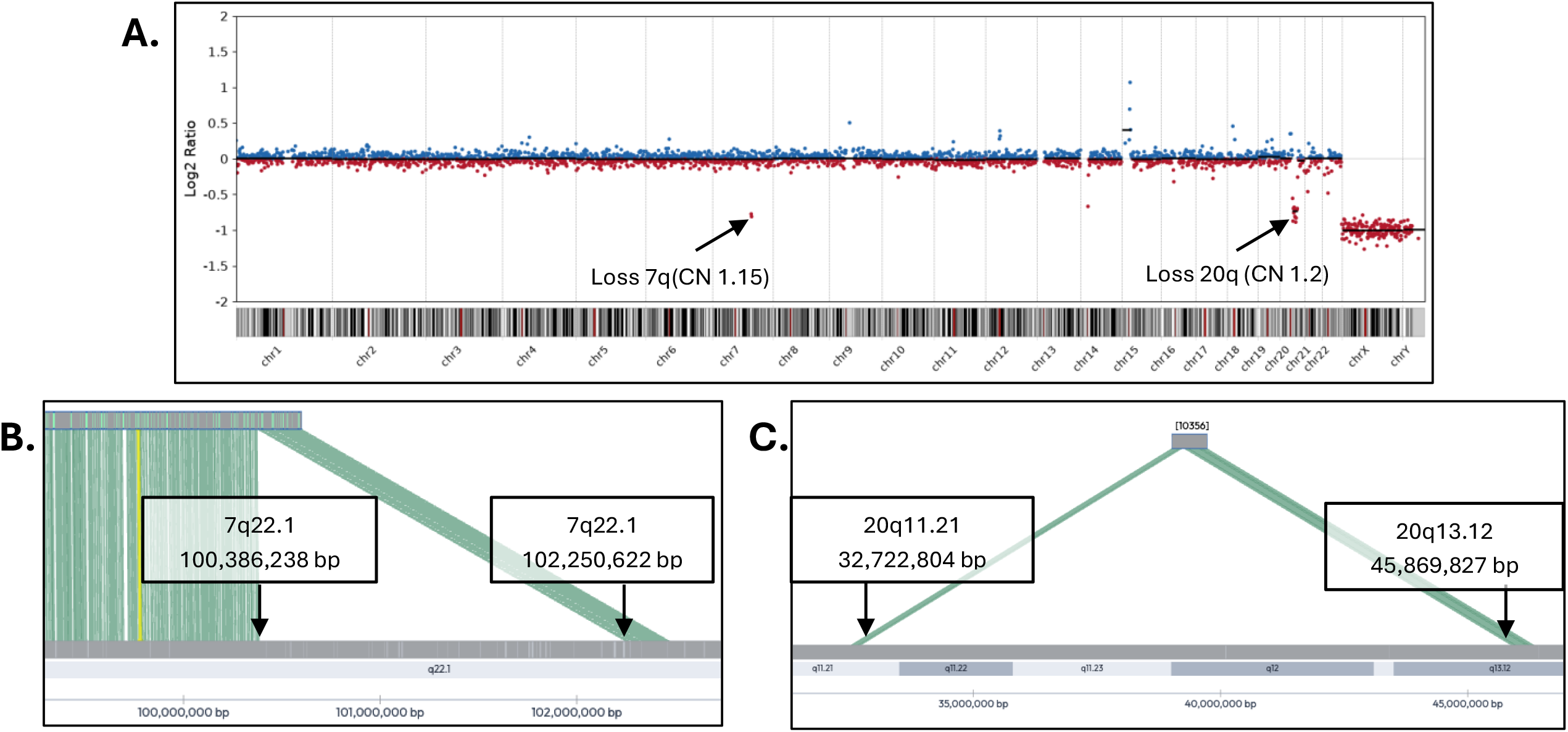
Copy number and structural variant characterization of interstitial losses in AUG-664. **(A)** The EGM copy number pipeline identified interstitial losses involving 7q22.1 (∼1.8Mb, estimated CN of 1.15) and 20q11.21q13.12 (∼13.1Mb, estimated CN of 1.20). Structural variant analysis defined the corresponding deletion breakpoints at **(B)** chr7:100,386,238–102,250,622 and (C) chr20:32,722,804– 45,869,827. Genomic coordinates are based on GRCh38.

**Figure 3.**
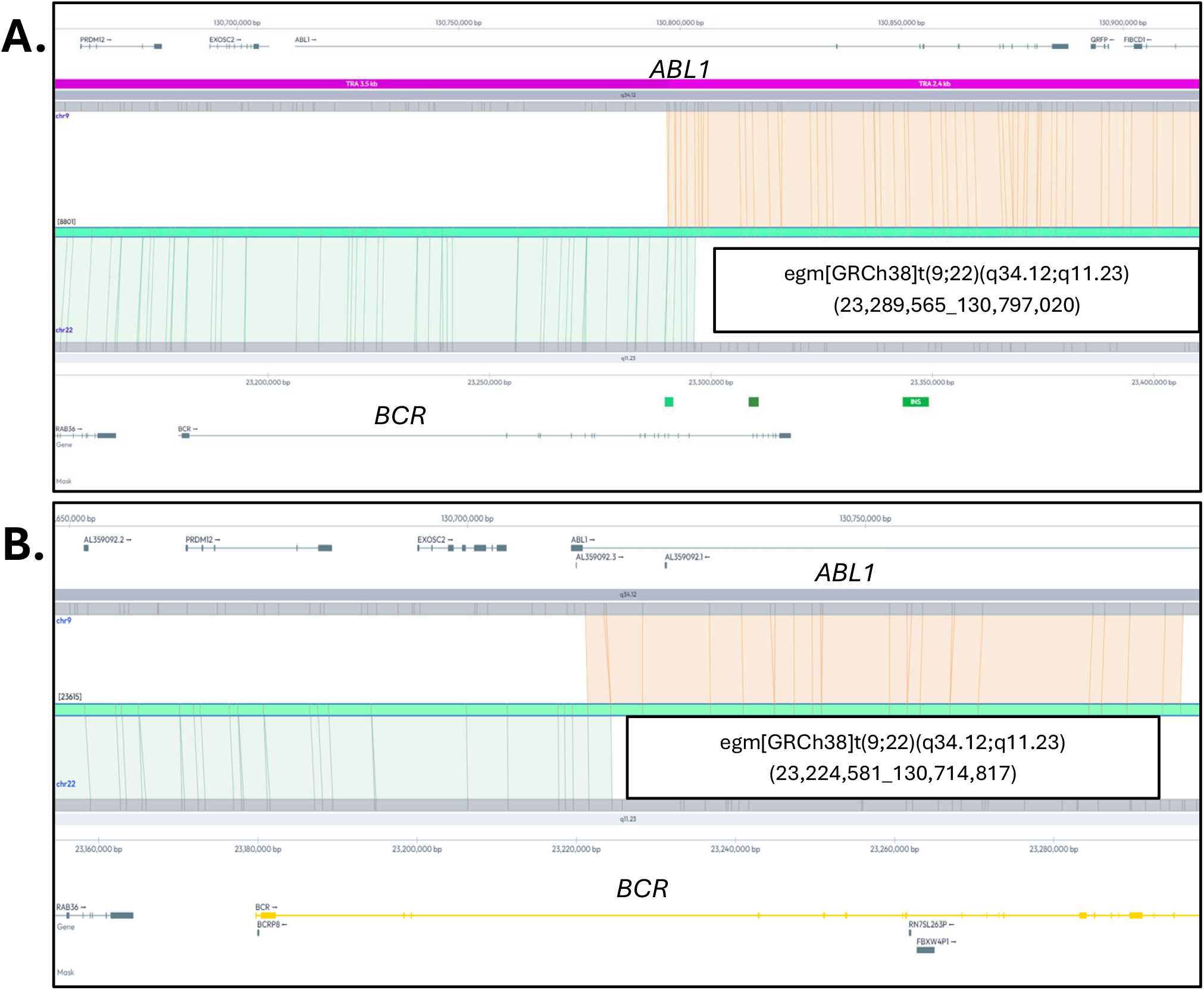
Breakpoint-level characterization of *BCR::ABL1* fusions by EGM. **(A)** AUG-679 and **(B)** AUG-667. Both translocation events were reported as t(9;22)(q34;q11.2) by karyotyping and as *BCR::ABL1*-positive by FISH. EGM better characterized the breakpoint positions in the two specimens as chr22:23,224,581 and chr9:130,714,817 in AUG-679, and chr22:23,289,565 and chr9:130,797,020 in AUG-667. Genomic coordinates are based on GRCh38.

## 4. Discussion

EGM uses electrical detection of high-molecular-weight (HMW) DNA molecules on the OhmX instrument to generate genome-wide contigs, with DNA labeling occurring at intervals of approximately 4 – 4.54 kb across the genome. Accurate detection and identification of pathogenic structural variants is important for clinical research diagnostics^1^. In this proof-of-principle pilot study, five bone marrow aspirate (BMA) specimens from two hematologic malignancy subtypes (AML and CML) were investigated for technology performance and evaluation and compared to chromosomal abnormalities reported by SOC techniques such as karyotyping, FISH and OGM.

Multiple studies from Augusta University have been published demonstrating the validation and performance of OGM for hematological malignancies and constitutional disorders^7,14,15^. This cohort included several distinct classes of structural variations: recurrent inter-chromosomal rearrangements, whole-chromosome gains and loss, segmental copy number loss, and an intra-chromosomal rearrangement. Most remarkably, EGM identified all previously reported SVs and CNVs. The 100% concordance observed across all SV classes provides an initial demonstration that EGM can reliably detect both structural and copy number abnormalities in myeloid malignancy specimens.

Genome mapping techniques have been recognized for the past several years for their potential to detect all SV classes at an unprecedented resolution in a single workflow^16,17^. Notably, an extensive body of OGM literature describing the application of long-range genome mapping in hematologic malignancy specimens has been published^6,7,10,11,12^. EGM addresses the same general analytical problem through an electronic single-molecule detection architecture. AUG-664 illustrates the complementary information generated by the EGM analysis pipelines. The copy number analysis pipeline identified the loss of 20q material, whereas the structural variant analysis pipeline described the junction underlying that loss as a fusion event, fus(20;20)(q11.21;q13.12). These are complementary representations of a single rearrangement rather than independent abnormalities. Additionally, EGM and OGM also identified the *KMT2A-PTD* variant as an insertion and a ∼1.8Mb copy number loss of 7q22.1 involving 45 genes, of which only *CUX1* is associated with cancer that was not detected by KT or FISH, demonstrating how genome-wide mapping techniques can provide high resolution structural and copy number information.

The scope of this pilot study was focused on technical concordance in a small, previously characterized set of hematological malignancy specimens. The cohort was enriched for known pathogenic abnormalities including 2 samples with the *BCR::ABL1* fusion events (AUG-679 and AUG-667). Importantly, Figure 3 demonstrates how EGM better characterizes the breakpoint resolution of the t(9;22) event. Additional studies on hematologic malignancies of various subtypes and larger sample sizes can build on these findings by assessing additional variant classes, different variant allele fractions, specimen characteristics, and complex genomes.

Taken together, the observed 100% concordance of EGM with standard-of-care methods demonstrates the ability of EGM to be a valuable tool for genome-wide assessment of structural and copy number variations in previously characterized AML and CML specimens.

## 5. Conclusions

In this proof-of-principle study, EGM demonstrated complete concordance with the standard-of-care cytogenetic abnormalities and OGM findings represented in five previously characterized myeloid malignancy specimens. Beyond concordance, EGM provided genome-wide structural and copy number characterization and refined the architecture and breakpoints of selected abnormalities. These findings establish technical feasibility and support larger prospective studies across diverse hematologic malignancies to define analytical performance, clinical utility, workflow characteristics, and the potential role of EGM as a first-line cytogenomic testing strategy.

## 8. Data Availability Statement

All the data produced in the present study are available upon reasonable request to the authors.

## 9. Acknowledgements

We would like to acknowledge Anna Lozar, Hanae Sugiura, Sayaka Tanaka, Lily Nasanovsky, Eiichi Araki, Mel Davey, and Scott Collins for their support during this study.

